# Peripheral immunoinflammatory markers and clinical correlations in Parkinson’s disease patients carrying *LRRK2* G2385R and R1628P variants

**DOI:** 10.64898/2026.09.20.26363505

**Authors:** Hans Xing Ding, Tzi Shin Toh, Nur Jannah Zulkefli, Nurul Syuhada Zulhaimi, Anis Nadhirah Khairul Anuar, Jia Wei Hor, Yi Wen Tay, Ing Xian Kong, Yong Chiang Pang, Reena Rajasuriar, Shen-Yang Lim, Ai Huey Tan, Lei Cheng Lit

## Abstract

Peripheral inflammation in Parkinson’s disease (PD) remains poorly characterised among carriers of Asian-prevalent *LRRK2* G2385R and R1628P variants. We investigated plasma immunoinflammatory markers (i.e., IL-6, TNF-α, CCL2, CX3CL1, CCL5, and VCAM-1) via multiplex immunoassay in 240 participants: PD-G2385R (n=53), PD-R1628P (n=58), PD-G2385R+R1628P (n=5), idiopathic PD (iPD; n=62), and controls (n=62), and their clinicobiological correlates including monocyte LRRK2 kinase activity (pRab10^Thr73^). Compared with iPD and/or controls, TNF-α, CCL2, CX3CL1, and CCL5 were lower in PD-G2385R, while IL-6, TNF-α, CCL2, and CCL5 were lower in PD-R1628P. Higher LRRK2 kinase activity correlated with lower TNF-α. CX3CL1 correlated with greater motor severity in iPD but less disability in both variant groups. CCL5 correlated with less motor complications in PD-G2385R, while IL-6 and VCAM-1 correlated with worse disability and/or cognition in PD-R1628P. These findings suggest that immunoinflammatory profiles and their clinical relevance in PD differ by genotype, with implications for stratification in future immunomodulatory trials.

## Introduction

The prevalence of Parkinson’s disease (PD) is fast increasing, including in Asian countries with large populations, and the disease is estimated to affect ∼25 million people by 2050^1–3^. Over the past few decades, advances in genetic research have significantly deepened our understanding of PD pathogenesis, identifying *Leucine Rich Repeat Kinase (LRRK2)* mutations as the most prevalent cause of familial forms of the disease^4,5^. There is distinct geographical variability in *LRRK2* variants, with the *LRRK2* G2019S variant predominating in Europeans and representing the most studied variant, but it is rare or absent in Asians^2,4,5^. In contrast, *LRRK2* G2385R and R1628P are prevalent in Asia populations, but remain comparatively understudied^2,5,6^.

Peripheral inflammation is one of the key pathophysiological features of PD^7,8^, supported by evidence of elevated levels of circulating proinflammatory cytokines and chemokines (i.e., IL-6, TNF-α, IL-1β, CRP, CCL2)^9,10^, along with a reduction in anti-inflammatory cytokines (i.e., IL-4)^10^, and alterations in the composition of peripheral immune cells^11^. Importantly, in PD, peripheral inflammatory markers have been linked to both the severity of motor and non-motor symptoms^10^ and longitudinal motor and non-motor progression^12–14^.

Studies of peripheral inflammation in PD associated with *LRRK2* variants remain relatively limited. Among the *LRRK2* variants, G2019S has been more extensively studied and is characterised by hyperactivity of LRRK2 kinase activity via increased phosphorylation of Rab10 at Thr73^15,16^. This variant has been shown in preclinical studies to drive overactivation of NF-κB and MAPK signalling pathways^17,18^, which are associated with the release of proinflammatory cytokines^19,20^. LRRK2 is also known to modulate monocytes in inflammatory responses by regulating endothelial adhesion, chemotaxis, and subsequent downstream signalling events^18,20–22^. However, studies on peripheral inflammatory markers in LRRK2 variant carriers remain scarce. Existing evidence, derived largely from the LRRK2 Cohort Consortium (LCC), has reported increased levels of chemokine CCL2 and/or growth factor PDGF in G2019S carriers compared to iPD^23,24^ and various additional markers when comparing *LRRK2* variant carriers (predominantly G2019S and R1441G/C/H carriers) to non-carriers irrespective of disease status^25^.

It is unclear whether PD associated with Asian-prevalent *LRRK2* variants (G2385R and/or R1628P), which confer milder but functionally-relevant increases in kinase activity^15,26^, follows a similar peripheral immunoinflammatory profile as G2019S. This study therefore aimed to characterise a range of key proinflammatory cytokines (IL-1β, IL-6, TNF-α), chemokines (CCL2, CX3CL1, CCL5), and the adhesion molecule VCAM-1 in the plasma of PD patients carrying *LRRK2* G2385R and/or R1628P variants, compared to idiopathic PD (iPD) and healthy controls, and to examine their associations with LRRK2 kinase activity and key clinical variables in PD.

## Methods

### Participant recruitment

PD patients were recruited from the University of Malaya Medical Centre (UMMC) Neurology Clinics, based on their previously genotyped profiles for *LRRK2* G2385R and R1628P^6^. PD diagnosis was assigned by movement disorder neurologists based on standard clinical diagnostic criteria^27^. Age-matched healthy controls were patients’ spouses who were free of neurological disorders and tested negative for *LRRK2* G2385R and R1628P.

The exclusion criteria were: (1) vaccination in the past three months; (2) recent acute illness in the past one month; (3) history of autoimmune diseases; (4) use of chronic corticosteroid or other immune-modulating therapy in the last five years; and (5) comorbidities preventing reliable completion of study assessments. The study was approved by the UMMC Medical Research Ethics Committee (MECID No. 2022427-11195), and written informed consent was obtained from all participants.

### Clinical assessments

Demographic, medical, and body mass index (BMI) data were collected. PD severity was evaluated using the International Parkinson and Movement Disorder Society-Unified PD Rating Scale (MDS-UPDRS) (with motor assessments conducted during the patients’ “ON” medication state) as well as the Clinical Impression of Severity Index for PD (CISI-PD). Cognition was assessed using the Montreal Cognitive Assessment (MoCA).

### Multiplex immunoassay for plasma cytokines and chemokines

Peripheral venous blood was collected in EDTA tubes and centrifuged within two hours of collection. Each plasma sample was divided into aliquots and stored immediately at - 80°C. Plasma levels of proinflammatory cytokines (IL-1β, IL-6, TNF-α), chemokines (CCL2, CX3CL1, CCL5), and the adhesion molecule VCAM-1 were measured using Simple Plex multiplex cartridge kit for the ELLA system (BioTechne, USA) according to the manufacturer’s instructions.

### Multiplex quantitative immunoblotting for LRRK2 kinase activity

Monocyte isolates were collected from the participants and multiplex quantitative immunoblotting was performed for pRab10^Thr73,^ pLRRK2^Ser935^, total Rab10, total LRRK2, and GAPDH, as previously described^26^.

### Statistical analysis

Data were analysed using IBM SPSS Statistics for Macintosh, version 29.0.1.0. The Shapiro-Wilk test was used to assess normality. Between-group differences were examined using analysis of variance (ANOVA), Kruskal-Wallis, or chi-square tests, as appropriate. Pairwise comparisons of immunoinflammatory marker levels were conducted using the Dunn’s post hoc test, while Quade’s non-parametric ANCOVA was used to adjust for age and sex. Partial Spearman correlation was applied to evaluate the associations between immunoinflammatory marker levels with LRRK2 kinase activity and clinical variables, controlling for age and disease duration, except for CISI-PD item 3 (motor complications) and MDS-UPDRS Part IV, where adjustments were made for age at diagnosis (given the known association between younger age at PD onset and the occurrence/severity of motor response complications^28^) and disease duration. Patients receiving apomorphine therapy or deep brain stimulation (DBS) were excluded from analyses involving MDS-UPDRS and CISI-PD scores, except for CISI-PD item 4 (cognitive status).

### Results Study cohort

Table 1 summarises the clinico-demographic data of participants. A total of 240 participants were recruited, including iPD (n=62), PD-G2385R (n=53), PD-R1628P (n=58), double variant PD-G2385R+R1628P (n=5), as well as healthy controls (n=62). There were no significant differences between iPD, *LRRK2*-PD, and healthy controls in age, sex, BMI, and comorbidities; and no difference between iPD and *LRRK2*-PD in LEDD, or in the proportions of patients receiving deep brain stimulation or apomorphine therapy. Double-variant carriers had significantly worse motor severity and cognitive impairment, as reflected by higher MDS-UPDRS Part III and CISI-PD total scores, and lower MoCA scores, compared to the other PD subgroups.

**Table 1.**
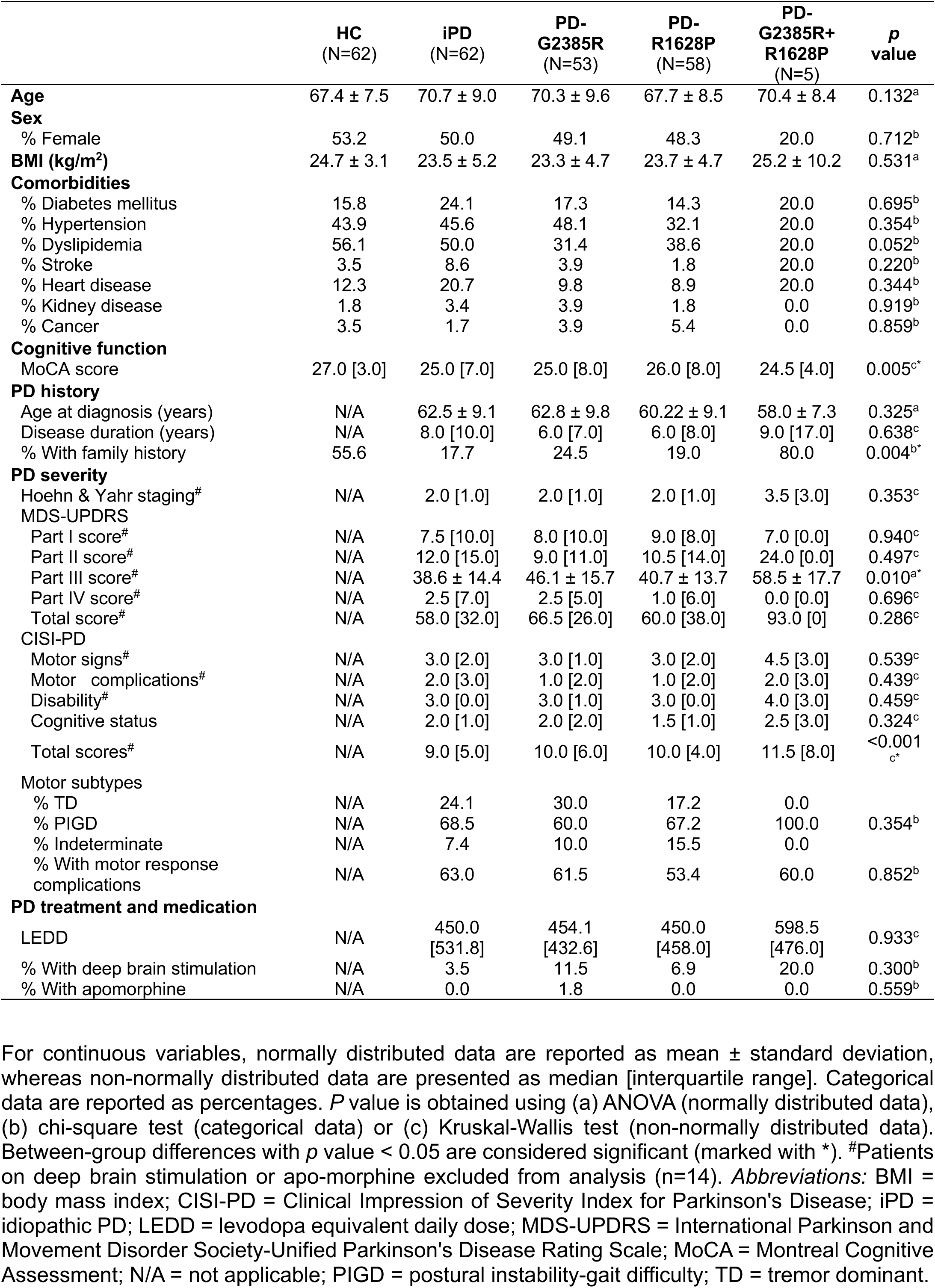
Demographic and clinical characteristics of the participants included in this study.

### Immunoinflammatory markers in patients and controls

Of the seven immunoinflammatory markers, IL-1β was below the lower limit of quantification (LLOQ <0.40 pg/mL) in most of the samples, despite the high and low internal controls falling within the expected ranges and repeated testing being performed. Hence, IL-1β was not considered further in our analysis. In the overall cohort (Figure 1), we found significant between-group differences in TNF-α (*p*<0.001), CCL2 (*p*=0.043), CX3CL1 (*p*=0.044), and CCL5 (*p*=0.018), but not in IL-6 (*p*=0.084) and VCAM-1 (*p*=0.925).

**Figure 1.**
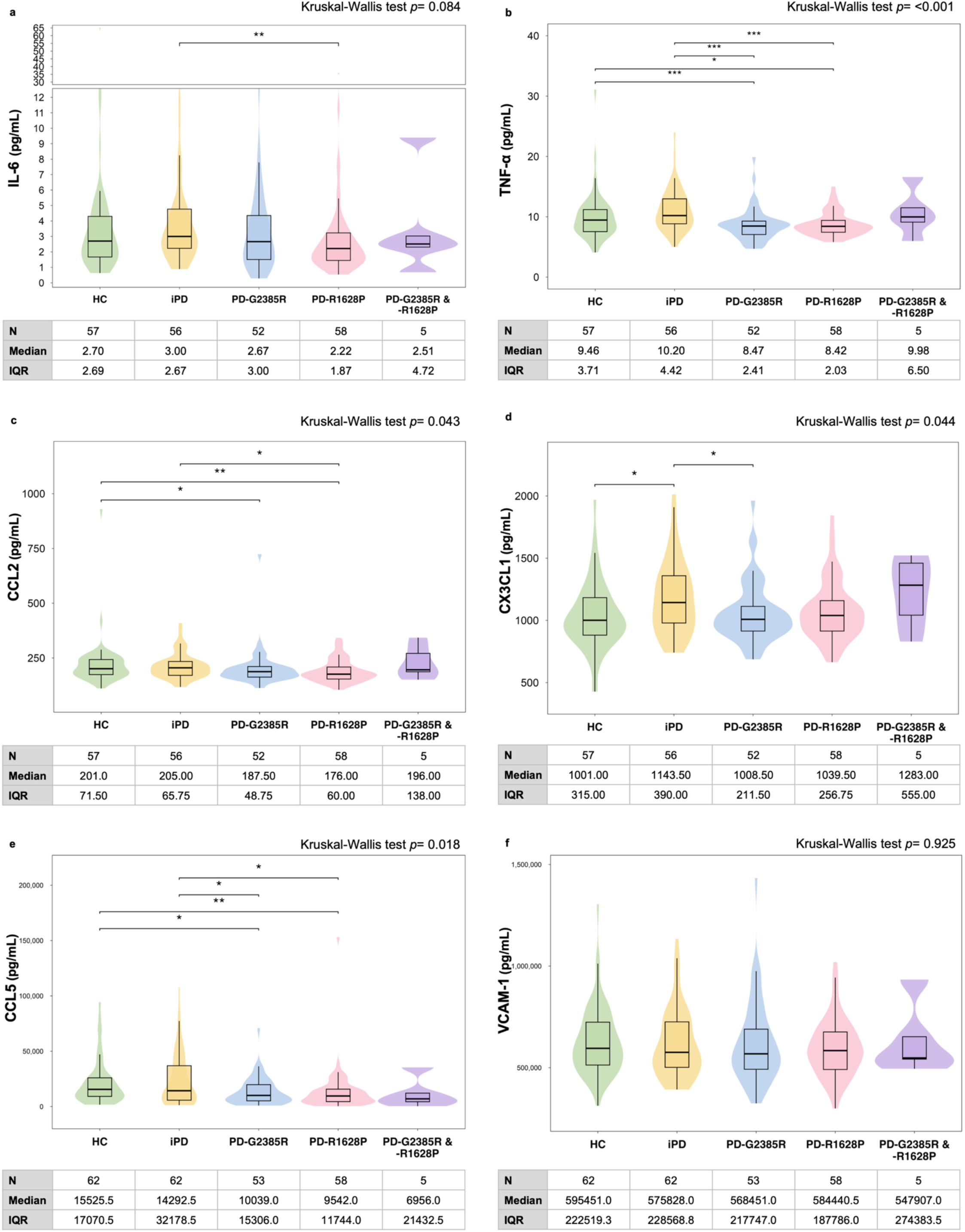
Comparison of immunoinflammatory marker levels across healthy controls, idiopathic PD (iPD), and *LRRK2*-PD groups. Box and violin plots show the distribution (median, minimum, and maximum) of immunoinflammatory marker levels for (a) IL-6, (b) TNF-α, (c) CCL2, (d) CX3CL1, (e) CCL5, and (f) VCAM-1 across five groups: HC (green), iPD (yellow), PD-G2385R (blue), PD-R1628P (pink) and PD-G2385R & R1628P (purple). Data were analysed using Quade’s non-parametric ANCOVA, adjusting for age and sex. Asterisks (*) indicate statistical significance: \**p* < 0.05, \*\**p* < 0.01, \*\*\**p* < 0.001. Only significant comparisons are shown, and pairwise comparisons using the Kruskal-Wallis test with post-hoc Dunn’s test are provided in the Supplementary Information. (g) Heatmap shows immunoinflammatory marker profiles across groups. Colour intensity reflects the median value of each immunoinflammatory marker, with blue denoting the highest median value and red the lowest across groups. Statistical significance is indicated as follows: Asterisk (*), *p* <0.05 versus HC; Hash (#), *p* <0.05 versus iPD.

In the post hoc analyses (Supplementary Tables 1 and 2), PD-G2385R patients had significantly lower TNF-α and CCL5 vs. healthy controls, and lower TNF-α and CX3CL1 vs. iPD. PD-R1628P patients had significantly lower TNF-α, CCL2, and CCL5 vs. healthy controls, and lower IL-6, TNF-α, CCL2, and CCL5 vs. iPD. These differences remained significant after adjusting for age and sex (Figure 1). Following covariate adjustment, we additionally observed a significant reduction in CCL2 (vs. healthy controls) and CCL5 (vs. iPD) among PD-G2385R patients. No significant differences were detected in double-variant *LRRK2* carriers compared to controls or iPD, nor among *LRRK2*-PD in any of the immunoinflammatory markers (Supplementary Table 3).

Meanwhile, iPD patients had increased TNF-α and CX3CL1 levels vs. healthy controls, however, the difference in TNF-α between iPD and controls was not significant after adjusting for age and sex.

### Correlations between immunoinflammatory markers and LRRK2 kinase activity

The present cohort was previously characterised for LRRK2 kinase activity^26^; here, we examined the correlation between LRRK2 kinase activity and plasma immunoinflammatory markers within the same participants. In the overall PD group, the ratio of pRab10^Thr73^/total Rab10 correlated negatively with TNF-α (r_s_=-0.225, *p*=0.003), which remained significant after adjusting for age and disease duration (r_s_=-0.170, *p*=0.034) (Figure 2a, Supplementary Table 4). The overall PD group was further stratified based on pRab10^Thr73^/total Rab10, where levels above the median for healthy controls (median=1.796) were considered to be high pRab10^Thr73^ phosphorylation. We found that the high pRab10^Thr73^ phosphorylation group had lower TNF-α, CCL2, and CX3CL1 levels, but only TNF-α remained significant after adjusting for age and disease duration (Figure 2b, Supplementary Table 5). In the overall PD group, no significant correlation was observed between any immunoinflammatory marker and the ratio of pLRRK2^Ser935^ /total LRRK2 (Supplementary Table 4).

**Figure 2.**
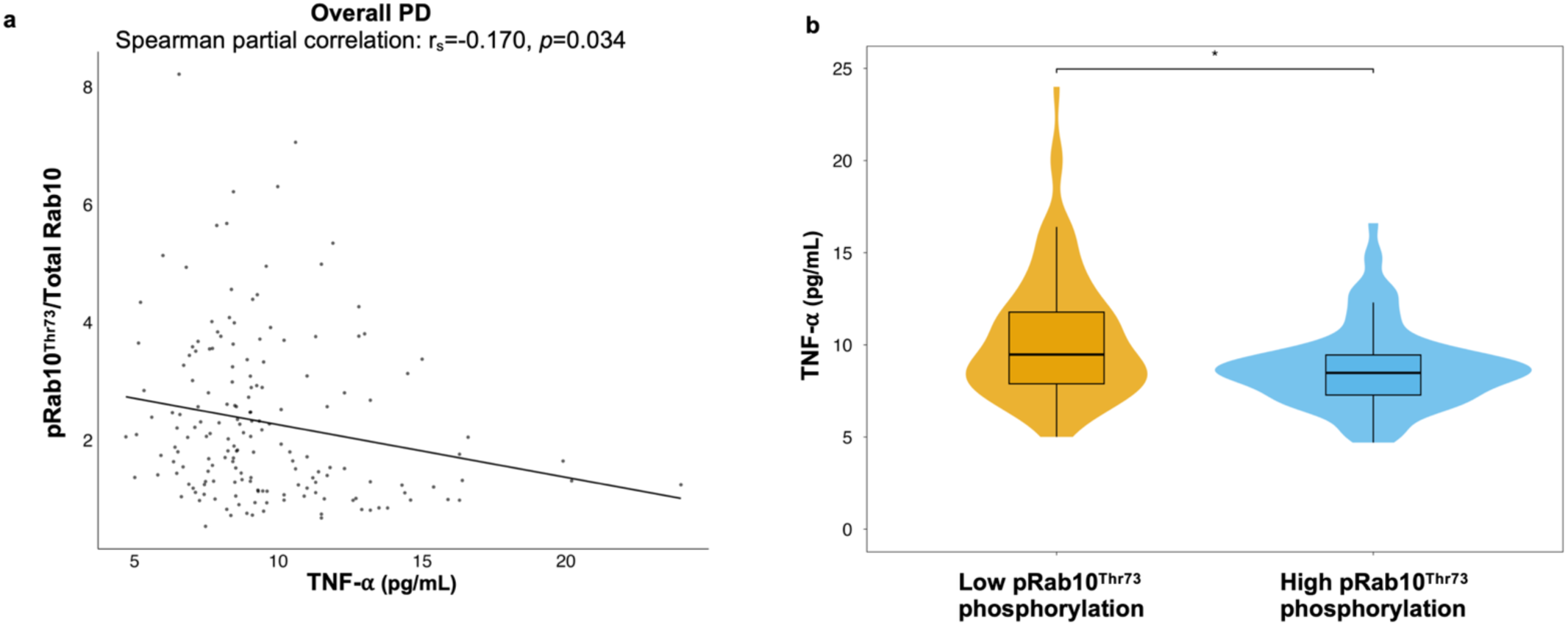
Correlation between immunoinflammatory markers and LRRK2 kinase activity in monocytes of PD. Scatter plots with partial Spearman correlation analysis showing (a) TNF-α levels versus p pRab10^Thr73^ /total Rab10 ratio in monocytes, adjusted for age and disease duration in the overall PD cohort. Each dot represents an individual with PD. Correlation coefficients (r_s_) and *p* values are indicated in the figure. (b) TNF-α levels stratified by higher or lower pRab10^Thr73^ phosphorylation than the median of healthy controls (median=1.796). Asterisks (*) indicate statistical significance: \**p* < 0.05, \*\**p* < 0.01, \*\*\**p* < 0.001. Only significant comparisons are shown, and pairwise comparisons were analysed using Quade’s non-parametric ANCOVA, adjusting for age and disease duration. Only significant comparisons are shown, and pairwise comparisons using the Mann-Whitney U test and Quade’s non-parametric ANCOVA are provided in the Supplementary Information.

### Correlations between immunoinflammatory markers and key clinical variables in PD

After adjusting for potential covariates (i.e., age/age at diagnosis and disease duration), we found several significant correlations between immunoinflammatory markers and clinical variables (Figure 3, Supplementary Table 6).

**Figure 3.**
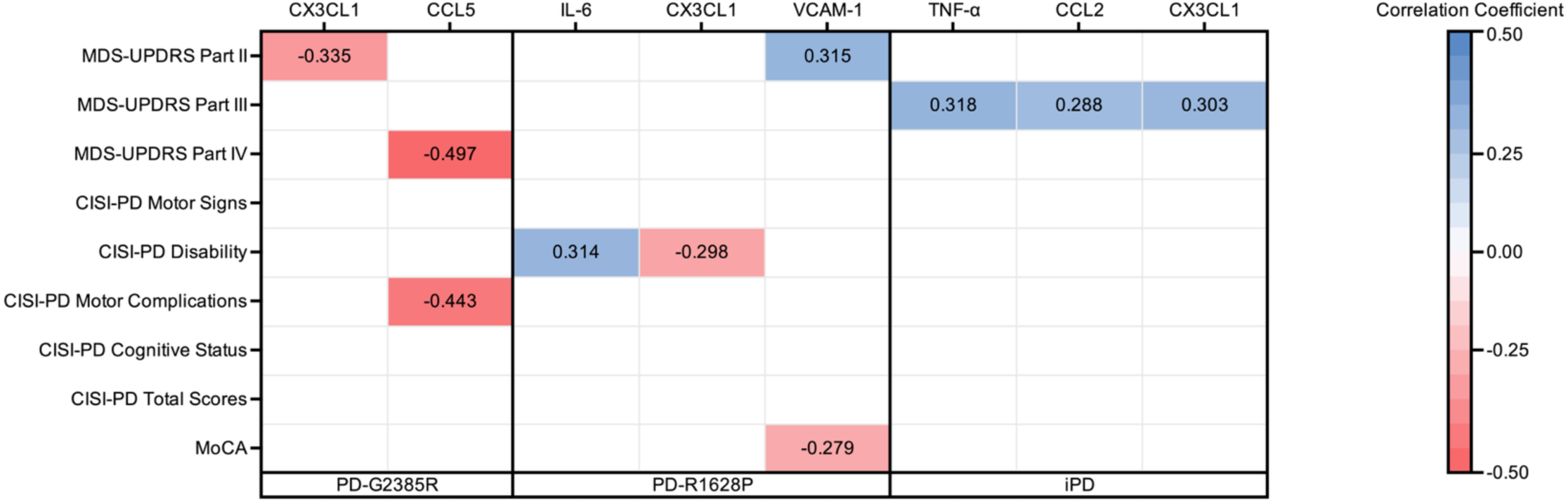
Clinical correlation heatmaps for individual immunoinflammatory markers. Heatmap of Spearman partial correlations between immunoinflammatory markers and clinical severity measures across PD-G2385R, PD-R1628P, and idiopathic PD (iPD). Correlations were adjusted for age and disease duration, except for analyses involving CISI-PD motor complications and MDS-UPDRS Part IV, which were adjusted for age at diagnosis and disease duration. Patients receiving apomorphine therapy or deep brain stimulation (DBS) were excluded from analyses involving MDS-UPDRS and CISI-PD scores, except for CISI-PD item 4 (cognitive status). Colour intensity represents correlation strength and direction (blue, positive; red, negative). Only statistically significant correlations (p < 0.05) are shown. Clinical measures include the MDS-Unified Parkinson’s Disease Rating Scale (MDS-UPDRS), Clinical Impression of Severity Index for Parkinson’s Disease (CISI-PD), and Montreal Cognitive Assessment (MoCA). Unadjusted correlations are provided in the Supplementary Information.

In the PD-G2385R subgroup, higher CX3CL1 levels correlated with less disability [MDS-UPDRS Part II (r_s_=-0.335, *p*=0.030)], whereas higher CCL5 levels correlated with less severe motor response complications [MDS-UPDRS Part IV (r_s_=-0.497; *p*=0.001); CISI-PD motor complications (r_s_=-0.443; *p*=0.003)].

Meanwhile, in the PD-R1628P subgroup, CISI-PD Disability scores correlated positively with IL-6 levels (r_s_=0.314; *p*=0.023) but inversely with CX3CL1 (r_s_=-0.298; *p*=0.032).

Higher VCAM-1 levels were associated with worse disability [MDS-UPDRS Part II (r_s_=0.315; *p*=0.024)] and poorer cognitive performance [MoCA (r_s_=-0.279; *p*=0.041)].

In iPD, higher TNF-α, CCL2, and CX3CL1 correlated with greater motor severity [MDS-UPDRS Part III (TNF-α: r_s_=0.318; *p*=0.028; CCL2: r_s_=0.288; *p*=0.047; CX3CL1: r_s_=0.303; *p*=0.036)].

## Discussion

Using a robust multiplex immunoassay, we identified a distinct peripheral immunoinflammatory profile in PD patients carrying Asian-prevalent *LRRK2* variants carriers compared to iPD and healthy controls. Both *LRRK2* G2385R and R1628P patients demonstrated significantly lower plasma proinflammatory cytokine and chemokine profiles (TNF-α, CCL2, CX3CL1, and CCL5 in PD-G2385R; IL-6, TNF-α, CCL2, and CCL5 in PD-R1628P) compared with iPD and healthy controls. Intriguingly, higher LRRK2 kinase activity correlated with lower plasma TNF-α levels across PD patients. Notably, these immunoinflammatory markers also demonstrated associations with clinically relevant measures of PD severity. Plasma CX3CL1 was associated with lower disability in both PD patient groups carrying *LRRK2* G2385R and R1628P, although it correlated with greater motor severity in iPD and was significantly higher in iPD vs. controls. Meanwhile, higher CCL5 was associated with less severe motor complications in PD-G2385R patients, whereas higher IL-6 and VCAM-1 levels were associated with greater disability and/or poorer cognition in PD-R1628P. Together, these findings suggest that immunoinflammatory profiles in PD differ by *LRRK2* genotype and may not follow a uniformly proinflammatory pattern. They provide biological insights into the clinical phenotype of *LRRK2*-assciated PD and, crucially, may be important factors to consider when selecting and stratifying PD patients for treatment with anti-inflammatory agents^7,29,30^.

In the present study, TNF-α, CCL2, CX3CL1, and CCL5 levels were lower in PD-G2385R while IL-6, TNF-α, CCL2, and CCL5 were lower in PD-R1628P, compared to iPD and/or HC. Our findings in PD patients with *LRRK2* G2385R and R1628P differ from prior reports involving PD patients with G2019S. Earlier work by Dzamko et al.^24^, which utilized a 27-marker panel including IL-6, TNF-α, CCL2, and CCL5, reported elevated IL-1β in non-manifesting *LRRK2* G2019S carriers compared to HC, but found no difference in other markers. Subsequently, Ahmadi Rastegar et al.^23^, using an overlapping panel on the same assay platform, further identified elevated CCL2 and PDGF in *LRRK2*-PD (predominantly *LRRK2* G2019S) compared to iPD, but found no difference in the remaining markers. More recently, Jaffery et al.^25^ used an expanded of 65 markers on the same assay platform, including IL-1β, IL-6, TNF-α, CX3CL1, and CCL2, and reported elevated CCL2 in *LRRK2* variant carriers, including both *LRRK2*-PD and non-manifesting *LRRK2* carriers, along with several other markers, but not IL-1β, IL-6, TNF-α, or CX3CL1. These analyses, conducted within overlapping LCC cohort samples, consistently implicated an increase in CCL2, but no difference in IL-6, TNF-α, CX3CL1, CCL5, all of which were examined in present study. Beyond the LCC cohort, the BEAT-PD study^31^ evaluated eight markers and similarly observed no significant differences in TNF-α, IL-1β, or IL-6 levels across LRRK2 variant carriers (both PD and non-manifesting), iPD, and healthy controls. In the present study, iPD showed increased CX3CL1 compared to HC, consistent with the report by Li et al.^32^

The lower levels of peripheral immunoinflammatory markers observed in PD patients carrying Asian-prevalent *LRRK2* variants may potentially be explained by several factors. The first relates to early life immune hyperactivation leading to immune exhaustion later in life. It has been postulated that *LRRK2* (as well as other PD gene) mutations may confer beneficial fitness effects in early life by increasing resilience to infection via, among other mechanisms, elevation of LRRK2 kinase activity leading to increased cytokine production^33^. However, sustained, unresolved inflammation increases PD risk later in life, and ultimately to immune exhaustion. This postulation is supported by recent preclinical work by Wallings et al.^35^, who examined the effect of LRRK2 kinase activity on myeloid immune function using young (2-3 months) and aged (18-21 months) *LRRK2* R1441C (gain-of-function) knock-in mice compared to age-matched wild-type control mice. In young R1441C mice, macrophages showed enhanced antigen presentation, pathogen clearance, and cytokine production compared to young wild-type control mice; in old age R1441C mice, this immune hyperactivation phenotype had collapsed into suppressed antigen presentation, hypophagocytosis, upregulated exhaustion marker expression (PD-L1), and decreased anti-inflammatory IL-10 production, compared to aged wild-type control mice. The same exhausted phenotype (hypophagocytosis, increased PD-L1) was also observed in macrophages from R1441C/Y1699C PD patients, but was absent in young, non-manifesting R1441C carriers. Another study found that peripheral IL-1β was elevated in non-manifesting G2019S carriers, but not in manifesting carriers^24^, again consistent with immune hyperactivation preceding disease onset and followed by immune exhaustion. Given the advanced age of our patients with *LRRK2* variants, the low levels of peripheral immunoinflammatory markers observed could reflect this immune exhausted state.

Secondly, the panel utilised in the present study, comprising IL-1β, IL-6, TNF-α CCL2, CX3CL1, CCL5, and VCAM-1, may not fully capture the alterations in immunoinflammatory profile in our patients. The present study’s focus on these markers was motivated by our interest in their role within monocyte-associated inflammatory pathways. However, LRRK2 is highly expressed in multiple immune cell populations^36^; thus, expanding the analysis to incorporate T cell and B cell markers, as well as anti-inflammatory markers, growth factors, and soluble receptors^25^, may provide more comprehensive insights into the immune states of PD patients carrying *LRRK2* G2385R and R1628P variants. An additional consideration is that peripheral markers alone may not be sufficient to capture LRRK2-associated immune activity in the central nervous system (CNS). Plasma inflammatory markers are imperfect proxies for CNS neuroinflammation, because central and peripheral inflammatory responses are partly compartmentalized and independently regulated despite bidirectional immune crosstalk. Notably, studies of PD patients with G2019S patients did not identify a correlation between cerebrospinal fluid (CSF) and peripheral cytokine levels, neither in the BEAT-PD study (TNF-α and IL-6)^31^, nor in the LCC cohort (IL-6, CCL2, TNF-α, and CX3CL1)^25^. More broadly in iPD, studies examining the correlation between inflammatory markers in CSF and blood remain scarce, with the few available studies reporting no correlation between CSF and plasma levels of IL-6 and TNF-α^37,38^. Therefore, the lower levels of peripheral immunoinflammatory markers observed in the present study should not be interpreted as direct evidence of reduced neuroinflammation in Asian-prevalent *LRRK2*-PD patients. Further studies utilising paired sampling could provide a more direct measure of neuroinflammation, however, recognising that there are substantial challenges associated with CSF sampling in our local/regional context^39^.

In present study, higher pRab10^Thr73^ phosphorylation in monocytes, used as a measure of LRRK2 kinase activity^15^, was found to correlate with lower TNF-α levels across PD patients, but not with other immunoinflammatory markers in our study. In contrast, a prior study in iPD reported no significant association between TNF-α and pRab10^Thr73^ phosphorylation in neutrophils and peripheral blood mononuclear cells (PBMCs)^40^. Meanwhile, a preclinical study reported that LRRK2 kinase activity and inhibitor treatment did not have a major effect on Toll-like receptor-stimulated cytokine levels in iPSC and human-derived immune cells^41^. Taken together, the inverse association between peripheral TNF-α levels and LRRK2 kinase activity in PD patients requires further validation studies in independent cohorts. Notably, the modestly low immunoinflammatory profile in Asian-prevalent *LRRK2* variant carriers aligns with our previous observation of a milder clinical phenotype and slower disease progression in this cohort^6^. Our present study identified different association patterns between immunoinflammatory markers and clinical severity in each PD subgroup. This divergence suggests that the mechanisms linking peripheral immune dysregulation to clinical manifestations may differ across genotypes, and that cytokine profiles may not be generalised as universal biomarkers of disease pathogenesis or clinical severity in PD. Consistent with previous reports in iPD, we found that higher TNF-α and CCL2 correlated with greater disease severity^42^. The correlation between higher CCL2 levels and greater disease severity also aligns with LCC cohort observations comprising iPD and *LRRK2*-PD patients^23^.

This study has several limitations. First, the sample size of patients was moderate and, in particular, the number of double-variant *LRRK2* carriers was small, despite the major efforts and resources that have gone into building this genetically-defined biomarker cohort^26^. Second, the cross-sectional data presented in this study preclude assessment of longitudinal changes in peripheral immune profiles with disease progression; this is an area of ongoing work for us. Third, as discussed above, we focused primarily on monocyte-associated inflammatory pathways, and research into other immune cell populations and their pathways, as well as anti-inflammatory mechanisms, would also be of interest, including if feasible stimulation-dependent assays. Fourth, although analyses were adjusted for key covariates, there could be other unaccounted-for confounders such as lifestyle factors and comorbidities (i.e., smoking, diabetes, and cardiovascular disease), that may have influenced our outcome measures. Future studies utilising larger, longitudinally studied cohorts and comprehensive immunoprofiling may provide further insights into the role of LRRK2 in immune regulation and dysfunction in *LRRK2* variant carriers, as well as in PD more broadly.

## Conclusion

In conclusion, our study demonstrated an altered peripheral immunoinflammatory profile in Asian-prevalent *LRRK2* variant carriers. These findings suggest that immunoinflammatory mechanisms associated with these variants may differ from those at play in iPD. Further research is essential to better delineate the immune mechanisms modulating LRRK2-associated PD. An improved understanding of these peripheral immune mechanisms will be a step towards precision medicine implementation in the Asian PD population.

## Data Availability

All data produced in the present study are available upon reasonable request to the authors.

## Author contributions

A.H.T., S.Y.L., L.C.L., and R.R. contributed to the conception and design of the study. All authors contributed to data acquisition. H.X.D. and A.H.T. contributed to data analysis and interpretation of data. H.X.D. prepared the first draft of the manuscript. All authors reviewed, revised and approved the final manuscript.

## Acknowledgements

This work was supported by The Michael J. Fox Foundation, which supported a collaborative research effort between Universiti Malaya (Malaysia) and the University of Dundee (United Kingdom) for the discovery of Asian LRRK2 biomarkers (MJFF-010188, MJFF-021041, and MJFF-022659 awarded to A.H.T., S-.Y.L., and E.S.). The funder played no role in study design, data collection, analysis and interpretation of data, or the writing of this manuscript. We sincerely thank all participants for their invaluable contributions, without which this research would not have been possible.

## References

1. Dorsey, E. R., Sherer, T., Okun, M. S., & Bloem, B. R. (2018). The Emerging Evidence of the Parkinson Pandemic. Journal of Parkinson’s Disease, 8(Suppl 1), S3–S8. 10.3233/JPD-181474

2. Lim, S.-Y., Tan, A. H., Ahmad-Annuar, A., Klein, C., Tan, L. C. S., Rosales, R. L., Bhidayasiri, R., Wu, Y.-R., Shang, H.-F., Evans, A. H., Pal, P. K., Hattori, N., Tan, C. T., Jeon, B., Tan, E.-K., & Lang, A. E. (2019). Parkinson’s disease in the Western Pacific Region. The Lancet Neurology, 18(9), 865–879. 10.1016/S1474-4422(19)30195-4

3. Su, D., Cui, Y., He, C., Yin, P., Bai, R., Zhu, J., Lam, J. S. T., Zhang, J., Yan, R., Zheng, X., Wu, J., Zhao, D., Wang, A., Zhou, M., & Feng, T. (2025). Projections for prevalence of Parkinson’s disease and its driving factors in 195 countries and territories to 2050: Modelling study of Global Burden of Disease Study 2021. BMJ, 388, e080952. 10.1136/bmj-2024-080952

4. Lang, A. E., Hauser, R. A., Kalia, L. V., Hersh, B., Berger, Z., Arenas, R. L., Paisan-Ruiz, C., Fraser, K., Jennings, D., Kluss, J. H., Huntwork-Rodriguez, S., Henry, A. G., & Greenamyre, J. T. (2026). LRRK2 as a Potential Disease-Modifying Target in Sporadic Parkinson’s Disease. Movement Disorders, 41(2), 297–314. 10.1002/mds.70100

5. Lim, S.-Y., Tan, A. H., Ahmad-Annuar, A., Okubadejo, N. U., Lohmann, K., Morris, H. R., Toh, T. S., Tay, Y. W., Lange, L. M., Bandres-Ciga, S., Mata, I., Foo, J. N., Sammler, E., Ooi, J. C. E., Noyce, A. J., Bahr, N., Luo, W., Ojha, R., Singleton, A. B., … Klein, C. (2024). Uncovering the genetic basis of Parkinson’s disease globally: From discoveries to the clinic. The Lancet Neurology, 23(12), 1267–1280. 10.1016/S1474-4422(24)00378-8

6. Goh, Lim, J. L., Toh, T. S., Ong, R. Y., Yong, Q. H., Lew, C. C. Y., Hor, J. W., Tay, Y. W., Zulkefli, J., Khairul Anuar, A. N., Ding, H. X., Schee, J. P., Beh, Y. Y., Ibrahim, K. A., Mawardi, A. S., Lim, T. T., Looi, I., Chia, Y. K., Ooi, J. C. E., … Tan, A. H. (2025). LRRK2 p.G2385R and p.R1628P variants in a multi-ethnic Asian Parkinson’s Cohort: Epidemiology and clinical insights. Npj Parkinson’s Disease, 11(1), 320. 10.1038/s41531-025-01166-x

7. Tansey, M. G., Wallings, R. L., Houser, M. C., Herrick, M. K., Keating, C. E., & Joers, V. (2022). Inflammation and immune dysfunction in Parkinson disease. Nature Reviews. Immunology, 22(11), 657–673. 10.1038/s41577-022-00684-6

8. Williams, G. P., Schonhoff, A. M., Sette, A., & Lindestam Arlehamn, C. S. (2022). Central and Peripheral Inflammation: Connecting the Immune Responses of Parkinson’s Disease. Journal of Parkinson’s Disease, 12(s1), S129–S136. 10.3233/JPD-223241

9. Qin, X.-Y., Zhang, S.-P., Cao, C., Loh, Y. P., & Cheng, Y. (2016). Aberrations in Peripheral Inflammatory Cytokine Levels in Parkinson Disease: A Systematic Review and Meta-analysis. JAMA Neurology, 73(11), 1316–1324. 10.1001/jamaneurol.2016.2742

10. Qu, Y., Li, J., Qin, Q., Wang, D., Zhao, J., An, K., Mao, Z., Min, Z., Xiong, Y., Li, J., & Xue, Z. (2023). A systematic review and meta-analysis of inflammatory biomarkers in Parkinson’s disease. NPJ Parkinson’s Disease, 9, 18. 10.1038/s41531-023-00449-5

11. Recinto, S. J., Posey, J. E. J., Lefter, N., Vitic, Z., Overgaard, M. Ø., Liu, L., Howden, A. J. M., Eyer, K., Romero-Ramos, M., Tansey, M. G., & Stratton, J. A. (2026). Profiling peripheral immune cells in Parkinson’s disease: A Scoping Review (p. 2026.02.17.706426). bioRxiv. 10.64898/2026.02.17.706426

12. Bartl, M., Dakna, M., Schade, S., Otte, B., Wicke, T., Lang, E., Starke, M., Ebentheuer, J., Weber, S., Toischer, K., Schnelle, M., Sixel-Döring, F., Trenkwalder, C., & Mollenhauer, B. (2023). Blood Markers of Inflammation, Neurodegeneration, and Cardiovascular Risk in Early Parkinson’s Disease. Movement Disorders, 38(1), 68–81. 10.1002/mds.29257

13. Kim, R., Kim, H.-J., Shin, J. H., Lee, C. Y., Jeon, S. H., & Jeon, B. (2022). Serum Inflammatory Markers and Progression of Nonmotor Symptoms in Early Parkinson’s Disease. Movement Disorders, 37(7), 1535–1541. 10.1002/mds.29056

14. Williams-Gray, C. H., Wijeyekoon, R., Yarnall, A. J., Lawson, R. A., Breen, D. P., Evans, J. R., Cummins, G. A., Duncan, G. W., Khoo, T. K., Burn, D. J., & Barker, R. A. (2016). Serum immune markers and disease progression in an incident Parkinson’s disease cohort (ICICLE-PD). Movement Disorders, 31(7), 995–1003. 10.1002/mds.26563

15. Kalogeropulou, A. F., Purlyte, E., Tonelli, F., Lange, S. M., Wightman, M., Prescott, A. R., Padmanabhan, S., Sammler, E., & Alessi, D. R. (2022). Impact of 100 LRRK2 variants linked to Parkinson’s disease on kinase activity and microtubule binding. Biochemical Journal, 479(17), 1759–1783. 10.1042/BCJ20220161

16. Steger, M., Tonelli, F., Ito, G., Davies, P., Trost, M., Vetter, M., Wachter, S., Lorentzen, E., Duddy, G., Wilson, S., Baptista, M. A., Fiske, B. K., Fell, M. J., Morrow, J. A., Reith, A. D., Alessi, D. R., & Mann, M. (2016). Phosphoproteomics reveals that Parkinson’s disease kinase LRRK2 regulates a subset of Rab GTPases. eLife, 5, e12813. 10.7554/eLife.12813

17. Chen, C.-Y., Weng, Y.-H., Chien, K.-Y., Lin, K.-J., Yeh, T.-H., Cheng, Y.-P., Lu, C.-S., & Wang, H.-L. (2012). (G2019S) LRRK2 activates MKK4-JNK pathway and causes degeneration of SN dopaminergic neurons in a transgenic mouse model of PD. Cell Death & Differentiation, 19(10), 1623–1633. 10.1038/cdd.2012.42

18. Hongge, L., Kexin, G., Xiaojie, M., Nian, X., & Jinsha, H. (2015). The role of LRRK2 in the regulation of monocyte adhesion to endothelial cells. Journal of Molecular Neuroscience: MN, 55(1), 233–239. 10.1007/s12031-014-0312-9

19. Guo, Q., Jin, Y., Chen, X., Ye, X., Shen, X., Lin, M., Zeng, C., Zhou, T., & Zhang, J. (2024). NF-κB in biology and targeted therapy: New insights and translational implications. Signal Transduction and Targeted Therapy, 9(1), 53. 10.1038/s41392-024-01757-9

20. Wallings, R. L., & Tansey, M. G. (2019). LRRK2 regulation of immune-pathways and inflammatory disease. Biochemical Society Transactions, 47(6), 1581–1595. 10.1042/BST20180463

21. Moehle, M. S., Daher, J. P. L., Hull, T. D., Boddu, R., Abdelmotilib, H. A., Mobley, J., Kannarkat, G. T., Tansey, M. G., & West, A. B. (2015). The G2019S LRRK2 mutation increases myeloid cell chemotactic responses and enhances LRRK2 binding to actin-regulatory proteins. Human Molecular Genetics, 24(15), 4250–4267. 10.1093/hmg/ddv157

22. Strader, S., & West, A. B. (2023). The interplay between monocytes, α-synuclein and LRRK2 in Parkinson’s disease. Biochemical Society Transactions, 51(2), 747–758. 10.1042/BST20201091

23. Ahmadi Rastegar, D., Ho, N., Halliday, G. M., & Dzamko, N. (2019). Parkinson’s progression prediction using machine learning and serum cytokines. Npj Parkinson’s Disease, 5(1), 14. 10.1038/s41531-019-0086-4

24. Dzamko, N., Rowe, D. B., & Halliday, G. M. (2016). Increased peripheral inflammation in asymptomatic leucine-rich repeat kinase 2 mutation carriers. Movement Disorders: Official Journal of the Movement Disorder Society, 31(6), 889–897. 10.1002/mds.26529

25. Jaffery, R., Zhao, Y., Ahmed, S., Schumacher, J. G., Ahn, J., Shi, L., Wang, Y., Tan, Y., Chen, K., Tawbi, H., Wang, J., Schwarzschild, M. A., Peng, W., & Chen, X. (2025). Soluble Immune Factor Profiles in Blood and CSF Associated with LRRK2 Mutations and Parkinson’s Disease. Npj Parkinson’s Disease, 365. 10.1038/s41531-025-01215-5

26. Toh, T. S., Lit, L. C., Lim, S.-Y., Hor, J. W., Lew, C. Y., Khairul Anuar, A. N., Tay, Y. W., Black, K., Lim, J. L., Zulkefli, J., Lim, K. S., Ding, H. X., Padmanabhan, S., Ahmad-Annuar, A., Tan, E. K., Alessi, D. R., Sammler, E., & Tan, A. H. (2026). Ex Vivo LRRK2 Activation in Asian G2385R and R1628P Variant Carriers and Idiopathic Parkinson’s Disease. Movement Disorders, n/a(n/a). 10.1002/mds.70314

27. Lees, A. J., Hardy, J., & Revesz, T. (2009). Parkinson’s disease. The Lancet, 373(9680), 2055–2066. 10.1016/S0140-6736(09)60492-X

28. Prange, S., Danaila, T., Laurencin, C., Caire, C., Metereau, E., Merle, H., Broussolle, E., Maucort-Boulch, D., & Thobois, S. (2019). Age and time course of long-term motor and nonmotor complications in Parkinson disease. Neurology, 92(2), e148–e160. 10.1212/WNL.0000000000006737

29. Lang, A. E., Hauser, R. A., Kalia, L. V., Hersh, B., Berger, Z., Arenas, R. L., Paisan-Ruiz, C., Fraser, K., Jennings, D., Kluss, J. H., Huntwork-Rodriguez, S., Henry, A. G., & Greenamyre, J. T. (2025). LRRK2 as a Potential Disease-Modifying Target in Sporadic Parkinson’s Disease. Movement Disorders: Official Journal of the Movement Disorder Society. 10.1002/mds.70100

30. Patel, B., Greenland, J. C., & Williams-Gray, C. H. (2024). Clinical Trial Highlights: Anti-Inflammatory and Immunomodulatory Agents. Journal of Parkinson’s Disease, 14(7), 1283–1300. 10.3233/JPD-240353

31. Thaler, A., Omer, N., Giladi, N., Gurevich, T., Bar-Shira, A., Gana-Weisz, M., Goldstein, O., Kestenbaum, M., Shirvan, J. C., Cedarbaum, J. M., Orr-Urtreger, A., Regev, K., Shenhar-Tsarfaty, S., & Mirelman, A. (2021). Mutations in GBA and LRRK2 Are Not Associated with Increased Inflammatory Markers. Journal of Parkinson’s Disease, 11(3), 1285–1296. 10.3233/JPD-212624

32. Li, Y., Yang, Y., Zhao, A., Luo, N., Niu, M., Kang, W., Xie, A., Lu, H., Chen, L., & Liu, J. (2022). Parkinson’s disease peripheral immune biomarker profile: A multicentre, cross-sectional and longitudinal study. Journal of Neuroinflammation, 19, 116. 10.1186/s12974-022-02481-3

33. Farrer, M. J. (2026). A unified evolutionary explanation of Parkinson’s disease. The Lancet Neurology, 25(9), 802–803. 10.1016/S1474-4422(26)00296-6

34. Alessi, D. R., & Sammler, E. (2018). LRRK2 kinase in Parkinson’s disease. Science, 360(6384), 36–37. 10.1126/science.aar5683

35. Wallings, R. L., McFarland, K., Staley, H. A., Neighbarger, N., Schaake, S., Brüggemann, N., Zittel, S., Usnich, T., Klein, C., Sammler, E. M., & Tansey, M. G. (2024). The R1441C-Lrrk2 mutation induces myeloid immune cell exhaustion in an age-and sex-dependent manner in mice. Science Translational Medicine, 16(772), eadl1535. 10.1126/scitranslmed.adl1535

36. Cook, D. A., Kannarkat, G. T., Cintron, A. F., Butkovich, L. M., Fraser, K. B., Chang, J., Grigoryan, N., Factor, S. A., West, A. B., Boss, J. M., & Tansey, M. G. (2017). LRRK2 levels in immune cells are increased in Parkinson’s disease. Npj Parkinson’s Disease, 3(1), 1–12. 10.1038/s41531-017-0010-8

37. Eidson, L. N., Kannarkat, G. T., Barnum, C. J., Chang, J., Chung, J., Caspell-Garcia, C., Taylor, P., Mollenhauer, B., Schlossmacher, M. G., Ereshefsky, L., Yen, M., Kopil, C., Frasier, M., Marek, K., Hertzberg, V. S., & Tansey, M. G. (2017). Candidate inflammatory biomarkers display unique relationships with alpha-synuclein and correlate with measures of disease severity in subjects with Parkinson’s disease. Journal of Neuroinflammation, 14, 164. 10.1186/s12974-017-0935-1

38. Wijeyekoon, R. S., Moore, S. F., Farrell, K., Breen, D. P., Barker, R. A., & Williams-Gray, C. H. (2020). Cerebrospinal Fluid Cytokines and Neurodegeneration-Associated Proteins in Parkinson’s Disease. Movement Disorders, 35(6), 1062–1066. 10.1002/mds.28015

39. Goh, Shambetova, C., Toh, T. S., Zhunusova, E., Tay, Y. W., Chua, J. P., Hann, W. H., Lim, J. Y. L., Tan, A. H., & Lim, S.-Y. (2026). Optimizing participant recruitment and retention in clinical studies by hearing the voice of persons with Parkinson’s. Journal of Parkinson’s Disease, 1877718X261479268. 10.1177/1877718X261479268

40. Atashrazm, F., Hammond, D., Perera, G., Bolliger, M. F., Matar, E., Halliday, G. M., Schüle, B., Lewis, S. J. G., Nichols, R. J., & Dzamko, N. (2019). LRRK2-mediated Rab10 phosphorylation in immune cells from Parkinson’s disease patients. Movement Disorders, 34(3), 406–415. 10.1002/mds.27601

41. Ahmadi Rastegar, D., Hughes, L. P., Perera, G., Keshiya, S., Zhong, S., Gao, J., Halliday, G. M., Schüle, B., & Dzamko, N. (2022). Effect of LRRK2 protein and activity on stimulated cytokines in human monocytes and macrophages. Npj Parkinson’s Disease, 8(1), 1–11. 10.1038/s41531-022-00297-9

42. Reale, M., Iarlori, C., Thomas, A., Gambi, D., Perfetti, B., Di Nicola, M., & Onofrj, M. (2009). Peripheral cytokines profile in Parkinson’s disease. Brain, Behavior, and Immunity, 23(1), 55–63. 10.1016/j.bbi.2008.07.003

