## Supplementary Materials for "Peripheral immunoinflammatory markers and clinical correlations in Parkinson’s disease patients carrying *LRRK2* G2385R and R1628P variants"

**Supplementary Table 1.** Comparison of immunoinflammatory marker levels with healthy controls.

| Comparison with healthy controls | *P* value | Adjusted *p* value |
|---|---|---|
| **IL-6** | | |
| iPD | 0.163 | 0.177 |
| PD-G2385R | 0.974 | 0.676 |
| PD-R1628P | 0.142 | 0.116 |
| PD-G2385R+R1628P | 0.920 | 0.908 |
| **TNF-α** | | |
| iPD | **0.026** | 0.080 |
| PD-G2385R | **0.027** | **<0.001** |
| PD-R1628P | **0.048** | **0.015** |
| PD-G2385R+R1628P | 0.551 | 0.819 |
| **CCL2** | | |
| iPD | 0.933 | 0.485 |
| PD-G2385R | 0.086 | **0.018** |
| PD-R1628P | **0.012** | **0.005** |
| PD-G2385R+R1628P | 0.742 | 0.899 |
| **CX3CL1** | | |
| iPD | **0.007** | **0.019** |
| PD-G2385R | 0.663 | 0.985 |
| PD-R1628P | 0.346 | 0.361 |
| PD-G2385R+R1628P | 0.119 | 0.138 |
| **CCL5** | | |
| iPD | 0.747 | 0.847 |
| PD-G2385R | **0.024** | **0.030** |
| PD-R1628P | **0.007** | **0.007** |
| PD-G2385R+R1628P | 0.167 | 0.210 |
| **VCAM-1** | | |
| iPD | 0.547 | 0.221 |
| PD-G2385R | 0.474 | 0.222 |

| | | |
|---|---|---|
| PD-R1628P | 0.389 | 0.328 |
| PD-G2385R+R1628P | 0.936 | 0.749 |

Pairwise comparisons between groups were conducted using the Kruskal-Wallis test with post-hoc Dunn's test. Age- and sex-adjusted *p* values were calculated using Quade's non-parametric ANCOVA. *P* values <0.05 (bold) were considered statistically significant.

**Supplementary Table 2.** Comparison of immunoinflammatory marker levels with iPD across *LRRK2*-stratified PD subgroups

| **Comparison with iPD** | ***P* value** | **Adjusted *p* value** |
|---|---|---|
| **IL-6** | | |
| PD-G2385R | 0.163 | 0.150 |
| PD-R1628P | **0.004** | **0.009** |
| PD-G2385R+R1628P | 0.507 | 0.590 |
| **TNF-α** | | |
| PD-G2385R | **<0.001** | **<0.001** |
| PD-R1628P | **<0.001** | **<0.001** |
| PD-G2385R+R1628P | 0.761 | 0.632 |
| **CCL2** | | |
| PD-G2385R | 0.104 | 0.091 |
| PD-R1628P | **0.016** | **0.038** |
| PD-G2385R+R1628P | 0.716 | 0.683 |
| **CX3CL1** | | |
| PD-G2385R | **0.027** | **0.023** |
| PD-R1628P | 0.075 | 0.146 |
| PD-G2385R+R1628P | 0.642 | 0.591 |
| **CCL5** | | |
| PD-G2385R | 0.052 | **0.047** |
| PD-R1628P | **0.017** | **0.013** |
| PD-G2385R+R1628P | 0.208 | 0.238 |
| **VCAM-1** | | |
| PD-G2385R | 0.890 | 0.964 |
| PD-R1628P | 0.787 | 0.821 |
| PD-G2385R+R1628P | 0.879 | 0.878 |

Pairwise comparisons between groups were conducted using the Kruskal-Wallis test with post-hoc Dunn's test. Age- and sex-adjusted *p* values were calculated using Quade's non-parametric ANCOVA. *P* values <0.05 (bold) were considered statistically significant.

**Supplementary Table 3.** Comparison of immunoinflammatory marker levels between PD-G2385R and PD-R1628P

| Comparison between PD-G2385R and PD-R1628P | *P* value | Adjusted *p* value |
|---|---|---|
| IL-6 | 0.161 | 0.264 |
| TNF-α | 0.779 | 0.379 |
| CCL2 | 0.463 | 0.734 |
| CX3CL1 | 0.628 | 0.383 |
| CCL5 | 0.705 | 0.653 |
| VCAM-1 | 0.902 | 0.793 |

Pairwise comparisons between groups were conducted using the Kruskal-Wallis test with post hoc Dunn's test. Age- and sex-adjusted *p* values were calculated using Quade's non-parametric ANCOVA. *P* values <0.05 (bold) were considered statistically significant.

**Supplementary Table 4.** Correlations of immunoinflammatory marker levels with pRab10$^{Thr73}$ and pLRRK2$^{Ser935}$ phosphorylation in the overall PD cohort.

| Correlation with pRab10$^{Thr73}$ and pLRRK2$^{Ser935}$ phosphorylation | Spearman's rank correlation coefficient | Spearman *p* value | Partial Spearman's rank correlation coefficient | Partial Spearman *p* value |
|---|---|---|---|---|
| **IL-6** | | | | |
| pRab10$^{Thr73}$ | -0.076 | 0.326 | -0.018 | 0.822 |
| pLRRK2$^{Ser935}$ | -0.057 | 0.481 | -0.065 | 0.418 |
| **TNF-α** | | | | |
| pRab10$^{Thr73}$ | **-0.225** | **0.003** | **-0.170** | **0.034** |
| pLRRK2$^{Ser935}$ | 0.069 | 0.390 | 0.079 | 0.331 |
| **CCL2** | | | | |
| pRab10$^{Thr73}$ | -0.122 | 0.112 | -0.033 | 0.688 |
| pLRRK2$^{Ser935}$ | 0.017 | 0.837 | 0.017 | 0.838 |
| **CX3CL1** | | | | |
| pRab10$^{Thr73}$ | -0.145 | 0.059 | -0.065 | 0.425 |
| pLRRK2$^{Ser935}$ | -0.021 | 0.795 | -0.024 | 0.767 |
| **CCL5** | | | | |
| pRab10$^{Thr73}$ | 0.072 | 0.350 | 0.008 | 0.925 |
| pLRRK2$^{Ser935}$ | -0.008 | 0.918 | -0.006 | 0.941 |

| VCAM-1 | | | | |
|---|---|---|---|---|
| pRab10$^{Thr73}$ | -0.015 | 0.844 | 0.072 | 0.375 |
| pLRRK2$^{Ser935}$ | -0.069 | 0.391 | -0.077 | 0.342 |

Spearman's correlations were shown between each immunoinflammatory marker and pRab10$^{Thr73}$ or pLRRK2$^{Ser935}$ phosphorylation in the overall PD cohort. Partial Spearman *p* values were adjusted for age and disease duration. Correlations with $p < 0.05$ are indicated in bold.

**Supplementary Table 5.** Comparison of immunoinflammatory marker levels between high pRab10$^{Thr73}$ and low pRab10 $^{Thr73}$ phosphorylation groups.

| Comparison between high pRab10$^{Thr73}$ and low pRab10$^{Thr73}$ phosphorylation | *P* value | Adjusted *p* value |
|---|---|---|
| IL-6 | 0.198 | 0.840 |
| TNF-α | **0.001** | **0.038** |
| CCL2 | **0.030** | 0.275 |
| CX3CL1 | **0.032** | 0.300 |
| CCL5 | 0.589 | 0.932 |
| VCAM1 | 0.538 | 0.567 |

The overall PD cohort was stratified into high pRab10$^{Thr73}$ and low pRab10$^{Thr73}$ phosphorylation, based on the median of pRab10$^{Thr73}$/total Rab10 ratio of healthy controls (median = 1.796). Those individuals with higher than 1.796 were grouped under the high pRab10$^{Thr73}$ phosphorylation group, whereas those with lower levels were grouped under the low pRab10$^{Thr73}$ phosphorylation group. Levels of immunoinflammatory markers were compared between the two groups using the Mann-Whitney U test and Quade's non-parametric analysis of covariance (ANCOVA), adjusting for age and disease duration. *P* values <0.05 are indicated in bold.

**Supplementary Table 6.** Correlations of immunoinflammatory marker levels with key clinical variables in *LRRK2*-stratified PD subgroups and iPD.

| PD-G2385R | | | | |
|---|---|---|---|---|
| Clinical Variables | Spearman's rank correlation | Spearman *p* value | Partial Spearman's correlation | Partial Spearman *p* value |
| **IL-6** | | | | |
| MDS UPDRS Part II | **0.356** | **0.016** | 0.161 | 0.307 |
| MDS UPDRS Part III | 0.143 | 0.348 | -0.033 | 0.836 |
| MDS-UPDRS Part IV | **-0.125** | **0.041** | -0.091 | 0.556 |

| | | | | |
|---|---|---|---|---|
| CISI-PD Motor Signs | 0.232 | 0.120 | -0.021 | 0.897 |
| CISI-PD Disability | 0.285 | 0.055 | 0.104 | 0.511 |
| CISI-PD Motor Complications | -0.089 | 0.558 | -0.045 | 0.774 |
| CISI-PD Cognitive Status | 0.259 | 0.064 | 0.003 | 0.981 |
| CISI-PD Total Scores | 0.216 | 0.150 | -0.041 | 0.797 |
| MoCA | **-0.282** | **0.045** | -0.088 | 0.547 |
| **TNF-α** | | | | |
| MDS UPDRS Part II | 0.153 | 0.317 | 0.003 | 0.985 |
| MDS UPDRS Part III | 0.054 | 0.723 | -0.074 | 0.642 |
| MDS-UPDRS Part IV | **-0.302** | **0.042** | -0.230 | 0.132 |
| CISI-PD Motor Signs | 0.190 | 0.205 | 0.002 | 0.992 |
| CISI-PD Disability | 0.150 | 0.319 | -0.008 | 0.958 |
| CISI-PD Motor Complications | -0.273 | 0.066 | -0.189 | 0.219 |
| CISI-PD Cognitive Status | 0.060 | 0.673 | -0.085 | 0.555 |
| CISI-PD Total Scores | 0.022 | 0.884 | -0.175 | 0.267 |
| MoCA | -0.272 | 0.054 | -0.180 | 0.216 |
| **CCL2** | | | | |
| MDS UPDRS Part II | 0.205 | 0.176 | 0.082 | 0.604 |
| MDS UPDRS Part III | 0.205 | 0.176 | 0.057 | 0.718 |
| MDS-UPDRS Part IV | -0.180 | 0.232 | -0.135 | 0.383 |
| CISI-PD Motor Signs | 0.278 | 0.062 | 0.137 | 0.388 |
| CISI-PD Disability | 0.232 | 0.121 | 0.102 | 0.520 |
| CISI-PD Motor Complications | -0.198 | 0.187 | -0.154 | 0.318 |
| CISI-PD Cognitive Status | 0.240 | 0.086 | 0.104 | 0.474 |
| CISI-PD Total Scores | 0.176 | 0.241 | 0.027 | 0.864 |
| MoCA | -0.143 | 0.318 | 0.021 | 0.888 |
| **CX3CL1** | | | | |
| MDS UPDRS Part II | -0.074 | 0.628 | **-0.335** | **0.030** |
| MDS UPDRS Part III | -0.059 | 0.698 | -0.201 | 0.203 |
| MDS-UPDRS Part IV | -0.082 | 0.586 | -0.023 | 0.884 |
| CISI-PD Motor Signs | 0.051 | 0.738 | -0.174 | 0.270 |
| CISI-PD Disability | 0.133 | 0.377 | -0.033 | 0.838 |
| CISI-PD Motor Complications | -0.155 | 0.304 | -0.104 | 0.502 |
| CISI-PD Cognitive Status | -0.053 | 0.707 | -0.218 | 0.128 |
| CISI-PD Total Scores | 0.022 | 0.887 | -0.199 | 0.207 |
| MoCA | -0.099 | 0.490 | 0.007 | 0.961 |
| **CCL5** | | | | |

| | | | | |
|---|---|---|---|---|
| MDS UPDRS Part II | -0.082 | 0.594 | 0.013 | 0.933 |
| MDS UPDRS Part III | 0.117 | 0.444 | 0.179 | 0.257 |
| MDS-UPDRS Part IV | **-0.502** | **<0.001** | **-0.497** | **0.001** |
| CISI-PD Motor Signs | -0.058 | 0.700 | -0.057 | 0.721 |
| CISI-PD Disability | -0.017 | 0.911 | 0.117 | 0.461 |
| CISI-PD Motor Complications | **-0.365** | **0.012** | **-0.443** | **0.003** |
| CISI-PD Cognitive Status | 0.035 | 0.804 | 0.096 | 0.507 |
| CISI-PD Total Scores | -0.177 | 0.234 | -0.168 | 0.288 |
| MoCA | 0.112 | 0.435 | 0.073 | 0.619 |
| **VCAM-1** | | | | |
| MDS UPDRS Part II | 0.108 | 0.481 | -0.209 | 0.185 |
| MDS UPDRS Part III | 0.072 | 0.639 | -0.151 | 0.339 |
| MDS-UPDRS Part IV | -0.143 | 0.342 | -0.065 | 0.675 |
| CISI-PD Motor Signs | 0.121 | 0.419 | -0.238 | 0.129 |
| CISI-PD Disability | 0.184 | 0.214 | -0.124 | 0.432 |
| CISI-PD Motor Complications | -0.184 | 0.215 | -0.124 | 0.423 |
| CISI-PD Cognitive Status | 0.175 | 0.209 | -0.079 | 0.587 |
| CISI-PD Total Scores | 0.128 | 0.392 | -0.211 | 0.180 |
| MoCA | **-0.311** | **0.026** | -0.137 | 0.348 |

| PD-R1628P | | | | |
|---|---|---|---|---|
| **Clinical Variables** | **Spearman's rank correlation** | **Spearman *p* value** | **Partial Spearman's correlation** | **Partial Spearman *p* value** |
| **IL-6** | | | | |
| MDS UPDRS Part II | 0.185 | 0.180 | 0.164 | 0.249 |
| MDS UPDRS Part III | **0.402** | **0.003** | 0.253 | 0.070 |
| MDS-UPDRS Part IV | -0.017 | 0.902 | 0.092 | 0.519 |
| CISI-PD Motor Signs | **0.319** | **0.019** | 0.148 | 0.294 |
| CISI-PD Disability | **0.398** | **0.003** | **0.314** | **0.023** |
| CISI-PD Motor Complications | 0.011 | 0.936 | 0.095 | 0.504 |
| CISI-PD Cognitive Status | 0.162 | 0.225 | 0.090 | 0.511 |
| CISI-PD Total Scores | 0.255 | 0.063 | 0.174 | 0.216 |
| MoCA | **-0.312** | **0.019** | -0.151 | 0.275 |
| **TNF-α** | | | | |
| MDS UPDRS Part II | -0.035 | 0.804 | -0.026 | 0.857 |
| MDS UPDRS Part III | **0.275** | **0.045** | 0.110 | 0.440 |

| | | | | |
|---|---|---|---|---|
| MDS-UPDRS Part IV | -0.209 | 0.129 | -0.043 | 0.764 |
| CISI-PD Motor Signs | 0.232 | 0.091 | 0.101 | 0.475 |
| CISI-PD Disability | 0.020 | 0.888 | -0.066 | 0.644 |
| CISI-PD Motor Complications | -0.129 | 0.351 | 0.025 | 0.859 |
| CISI-PD Cognitive Status | 0.128 | 0.339 | 0.112 | 0.411 |
| CISI-PD Total Scores | 0.045 | 0.746 | 0.016 | 0.909 |
| MoCA | **-0.276** | **0.040** | -0.164 | 0.237 |
| **CCL2** | | | | |
| MDS UPDRS Part II | -0.125 | 0.367 | -0.185 | 0.188 |
| MDS UPDRS Part III | -0.004 | 0.979 | -0.094 | 0.507 |
| MDS-UPDRS Part IV | -0.084 | 0.544 | -0.118 | 0.403 |
| CISI-PD Motor Signs | -0.028 | 0.838 | -0.146 | 0.301 |
| CISI-PD Disability | 0.021 | 0.880 | -0.073 | 0.608 |
| CISI-PD Motor Complications | -0.057 | 0.682 | -0.093 | 0.513 |
| CISI-PD Cognitive Status | 0.048 | 0.723 | -0.022 | 0.874 |
| CISI-PD Total Scores | -0.041 | 0.771 | -0.140 | 0.321 |
| MoCA | 0.051 | 0.706 | 0.136 | 0.328 |
| **CX3CL1** | | | | |
| MDS UPDRS Part II | -0.043 | 0.757 | -0.096 | 0.504 |
| MDS UPDRS Part III | -0.009 | 0.950 | -0.222 | 0.115 |
| MDS-UPDRS Part IV | 0.004 | 0.978 | 0.118 | 0.403 |
| CISI-PD Motor Signs | 0.034 | 0.809 | -0.173 | 0.220 |
| CISI-PD Disability | -0.129 | 0.353 | **-0.298** | **0.032** |
| CISI-PD Motor Complications | 0.071 | 0.608 | 0.170 | 0.228 |
| CISI-PD Cognitive Status | 0.112 | 0.401 | 0.052 | 0.705 |
| CISI-PD Total Scores | 0.044 | 0.752 | -0.067 | 0.636 |
| MoCA | -0.209 | 0.123 | -0.064 | 0.644 |
| **CCL5** | | | | |
| MDS UPDRS Part II | 0.026 | 0.850 | 0.047 | 0.741 |
| MDS UPDRS Part III | -0.015 | 0.916 | -0.003 | 0.981 |
| MDS-UPDRS Part IV | 0.106 | 0.448 | 0.114 | 0.422 |
| CISI-PD Motor Signs | 0.024 | 0.865 | 0.052 | 0.716 |
| CISI-PD Disability | 0.072 | 0.603 | 0.105 | 0.459 |
| CISI-PD Motor Complications | 0.171 | 0.216 | 0.190 | 0.177 |
| CISI-PD Cognitive Status | -0.083 | 0.535 | -0.083 | 0.545 |
| CISI-PD Total Scores | 0.042 | 0.762 | 0.077 | 0.590 |
| MoCA | 0.060 | 0.663 | 0.054 | 0.700 |

| | | | | |
|---|---|---|---|---|
| **VCAM-1** | | | | |
| MDS UPDRS Part II | **0.297** | **0.029** | **0.315** | **0.024** |
| MDS UPDRS Part III | 0.094 | 0.498 | -0.048 | 0.733 |
| MDS-UPDRS Part IV | -0.048 | 0.732 | 0.070 | 0.620 |
| CISI-PD Motor Signs | 0.141 | 0.310 | 0.022 | 0.875 |
| CISI-PD Disability | 0.087 | 0.531 | 0.017 | 0.906 |
| CISI-PD Motor Complications | -0.060 | 0.665 | 0.032 | 0.824 |
| CISI-PD Cognitive Status | 0.227 | 0.087 | 0.207 | 0.125 |
| CISI-PD Total Scores | 0.154 | 0.267 | 0.122 | 0.388 |
| MoCA | **-0.381** | **0.004** | **-0.279** | **0.041** |

| **iPD** | | | | |
|---|---|---|---|---|
| **Clinical Variables** | **Spearman's rank correlation** | **Spearman *p* value** | **Partial Spearman's correlation** | **Partial Spearman *p* value** |
| **IL-6** | | | | |
| MDS UPDRS Part II | 0.194 | 0.169 | 0.048 | 0.746 |
| MDS UPDRS Part III | 0.125 | 0.378 | 0.069 | 0.643 |
| MDS-UPDRS Part IV | -0.060 | 0.672 | -0.135 | 0.355 |
| CISI-PD Motor Signs | 0.229 | 0.102 | 0.115 | 0.437 |
| CISI-PD Disability | 0.164 | 0.244 | 0.060 | 0.685 |
| CISI-PD Motor Complications | 0.043 | 0.764 | 0.006 | 0.967 |
| CISI-PD Cognitive Status | 0.174 | 0.205 | 0.085 | 0.545 |
| CISI-PD Total Scores | 0.221 | 0.112 | 0.108 | 0.464 |
| MoCA | **-0.295** | **0.029** | -0.250 | 0.071 |
| **TNF-α** | | | | |
| MDS UPDRS Part II | **0.399** | **0.003** | 0.282 | 0.052 |
| MDS UPDRS Part III | **0.459** | **0.001** | **0.318** | **0.028** |
| MDS-UPDRS Part IV | -0.195 | 0.166 | -0.144 | 0.322 |
| CISI-PD Motor Signs | **0.332** | **0.016** | 0.103 | 0.484 |
| CISI-PD Disability | **0.305** | **0.028** | 0.141 | 0.339 |
| CISI-PD Motor Complications | -0.105 | 0.459 | -0.091 | 0.536 |
| CISI-PD Cognitive Status | 0.234 | 0.085 | -0.100 | 0.477 |
| CISI-PD Total Scores | 0.220 | 0.114 | 0.065 | 0.660 |
| MoCA | **-0.441** | **0.001** | -0.240 | 0.083 |
| **CCL2** | | | | |
| MDS UPDRS Part II | -0.031 | 0.829 | -0.213 | 0.146 |

| | | | | |
|---|---|---|---|---|
| MDS UPDRS Part III | **0.381** | **0.005** | **0.288** | **0.047** |
| MDS-UPDRS Part IV | **-0.281** | **0.043** | -0.236 | 0.102 |
| CISI-PD Motor Signs | 0.252 | 0.072 | 0.199 | 0.175 |
| CISI-PD Disability | 0.172 | 0.222 | 0.084 | 0.569 |
| CISI-PD Motor Complications | -0.205 | 0.146 | -0.198 | 0.173 |
| CISI-PD Cognitive Status | 0.120 | 0.381 | -0.093 | 0.507 |
| CISI-PD Total Scores | 0.013 | 0.925 | -0.111 | 0.453 |
| MoCA | -0.149 | 0.278 | 0.038 | 0.786 |
| **CX3CL1** | | | | |
| MDS UPDRS Part II | **0.320** | **0.021** | 0.105 | 0.477 |
| MDS UPDRS Part III | **0.428** | **0.002** | **0.303** | **0.036** |
| MDS-UPDRS Part IV | -0.045 | 0.752 | -0.143 | 0.327 |
| CISI-PD Motor Signs | **0.293** | **0.035** | 0.066 | 0.656 |
| CISI-PD Disability | 0.252 | 0.071 | 0.049 | 0.741 |
| CISI-PD Motor Complications | 0.079 | 0.577 | -0.019 | 0.899 |
| CISI-PD Cognitive Status | 0.264 | 0.051 | 0.060 | 0.667 |
| CISI-PD Total Scores | **0.308** | **0.025** | 0.088 | 0.551 |
| MoCA | **-0.277** | **0.041** | -0.101 | 0.470 |
| **CCL5** | | | | |
| MDS UPDRS Part II | **-0.295** | **0.034** | -0.145 | 0.327 |
| MDS UPDRS Part III | **-0.278** | **0.046** | -0.149 | 0.314 |
| MDS-UPDRS Part IV | -0.176 | 0.212 | -0.091 | 0.534 |
| CISI-PD Motor Signs | -0.238 | 0.087 | 0.048 | 0.745 |
| CISI-PD Disability | -0.250 | 0.071 | 0.012 | 0.934 |
| CISI-PD Motor Complications | -0.101 | 0.470 | 0.036 | 0.809 |
| CISI-PD Cognitive Status | -0.240 | 0.075 | -0.108 | 0.443 |
| CISI-PD Total Scores | -0.248 | 0.070 | -0.011 | 0.940 |
| MoCA | 0.106 | 0.427 | -0.026 | 0.852 |
| **VCAM-1** | | | | |
| MDS UPDRS Part II | 0.200 | 0.154 | -0.023 | 0.879 |
| MDS UPDRS Part III | 0.208 | 0.138 | 0.074 | 0.617 |
| MDS-UPDRS Part IV | -0.002 | 0.987 | -0.138 | 0.345 |
| CISI-PD Motor Signs | 0.228 | 0.101 | -0.084 | 0.570 |
| CISI-PD Disability | 0.201 | 0.149 | -0.077 | 0.605 |
| CISI-PD Motor Complications | 0.071 | 0.611 | -0.098 | 0.501 |
| CISI-PD Cognitive Status | **0.340** | **0.010** | 0.167 | 0.232 |
| CISI-PD Total Scores | 0.198 | 0.151 | -0.098 | 0.506 |

| | | | | |
|---|---|---|---|---|
| MoCA | -0.194 | 0.145 | -0.079 | 0.575 |

Partial Spearman correlations between immunoinflammatory markers and clinical severity variables across PD-G2385R, PD-R1628P, and idiopathic PD (iPD) were calculated with adjustment for age and disease duration, except for CISI-PD motor complications and MDS-UPDRS Part IV, which were adjusted for age at diagnosis and disease duration. Patients receiving apomorphine therapy or deep brain stimulation (DBS) were excluded from analyses involving MDS-UPDRS and CISI-PD scores, except for CISI-PD cognitive status. Unadjusted Spearman correlations are presented for comparison. Statistically significant correlations ($p < 0.05$) are indicated in bold.
